# Is calcium intake associated with sleep quality? : a systematic review

**DOI:** 10.1101/2024.12.03.24318236

**Authors:** Ami Isoda, Junko Kiriya, Masamine Jimba

## Abstract

As the prevalence of sleep disorders has grown worldwide, studies on sleep quality are increasingly required. Recent evidence suggests that calcium intake may influence sleep quality and duration, but few studies have investigated factors associated with sleep quality. This systematic review aimed to examine and synthesize existing evidence on the association between calcium intake and sleep quality and sleep duration. We included randomized controlled trials, case-control studies, cohort studies, and cross-sectional studies from multiple databases, including PubMed, Web of Science, Cochrane Library, Scopus, and CINAHL Plus, followed by keywords search at Google Scholar and reference review of included articles. A total of seven independent studies presented in nine reports met the inclusion criteria. Six of them were cross-sectional studies. The studies suggested a positive association between calcium intake and sleep quality, although the results were not statistically significant in some cases. Studies also indicated that lower calcium intake was associated with inappropriate sleep duration. We decided not to conduct meta-analysis because of the insufficient number of studies. The findings suggest a positive relationship between calcium intake and sleep quality. To test the effectiveness of calcium intake on sleep quality, a randomized controlled trial is required across different ethnicity and culture.

## 1. Introduction

Sleep disorders have become a global health concern, with wide-reaching consequences on daily functioning and long-term health outcomes. As for insomnia, for example, its prevalence is estimated to be as high as 10 to 60% of the total population of the world.^1, 2, 3^ Sleep is closely related to health, and it accounts for approximately as much as one-third of our whole life.^4^ According to the Centers for Disease Control and Prevention of the United States of America, lacking quantity or quality of sleep can cause health problems and badly affect our daily lives.^5^ In light of its increasing prevalence, sleep-wake disorder was added to the International Classification of Diseases 11th Revision (ICD-11) in 2018.^6^ This implies an urgent need for an improvement in sleep quality.

Recent evidence suggests that calcium intake may be a key factor in sleep quality. Those who live in countries that lack dietary calcium intake tend to have short sleep duration. This indicates a possible association between calcium intake and sleep. According to a survey by the Organisation for Economic Cooperation and Development (OECD), average sleep duration of Japanese people is the shortest among OECD countries, followed by South Korea.^7^ Another study on sleep duration in young adults 16-30 years old showed that people living in the Asian region had the shortest sleep duration compared to those in other regions.^8^ Although various countries were included in the Asian region in this study, if we look at the data from Japan and South Korea only, the difference in the sleep duration could be even bigger. Meanwhile, the distribution of calcium intake in Asian region including these two countries shows that the national average intake of calcium is very low, which is less than 600 mg per day.^9^

Results from basic research also show that calcium may play an important role in the regulation of sleep duration. Experiments using computational model and mice suggested an effect of calcium on sleep. Tatsuki et al. found that sleep duration increased or decreased in mice with modified genes related to the Ca(2+)-dependent hyperpolarization pathway.^10^ The same mechanism may be applicable to human.

As the prevalence of sleep disorders has increased worldwide, understanding the possible cause of those issues and coming up with solutions to them is fundamental. However, few studies have investigated factors associated with sleep quality. Therefore, this study aimed to explore and synthesize the existing evidence for the association of calcium intake with sleep quality as a potential solution to sleep problems.

## 2. Methods

### 2.1. Criteria and outcomes

We included randomized controlled trials, case-control studies, cohort studies, and cross-sectional studies. In addition to these study designs, we targeted studies with participants who were 18 years or older. Then we excluded studies that particularly focused on specific health conditions such as psychiatric disorders, pregnancy, and other conditions. We also excluded studies that focused on people under specific working conditions, such as rotating shifts, which can affect sleep quality and duration.

We set calcium intake (amount) from diet, including supplement, as an exposure variable. Sleep quality was set as the primary outcome, and sleep duration as the secondary outcome.

### 2.2. Identification of studies and data collection

#### Electronic searches and selection of studies

We searched online databases for relevant literature through five search systems (searched databases presented in brackets): PubMed (Medline and others), Web of Science (Medline), Cochrane Library (Cochrane Central Register of Controlled Trials), Scopus (full index), and EbscoHost (CINAHL Plus), using the search strategies presented in appendix 1. Thereafter, we conducted a keyword search (search terms are shown in appendix 1) in Google Scholar to minimize the risk of missing relevant articles. To thoroughly search for relevant articles, we also checked the titles and abstracts of the references of the reports identified in the main database search.

We included articles with titles and abstracts in English. We did not restrict the publication year of reports (up to the end of March 2024).

After the search, two reviewers independently screened the titles and abstracts of all records. Thereafter, we obtained the full texts of all potentially relevant records for further examination of eligibility, using the predefined inclusion and exclusion criteria for study design, participants, and exposure and outcome variables. The two reviewers read all the obtained reports and independently judged whether to include each report. Any disagreements between the two reviewers were resolved by discussion.

#### Data extraction and management

We extracted the following information and tabulated the data from the selected articles: authors’ names, publication year, language, title, study settings, country, study population, participants’ demographics and baseline characteristics, sample size, details of the exposure and control conditions, study design, recruitment and study completion rates, outcomes and times of measurement, assessment tool for the exposure variables, assessment tool for the outcome variables, effect measure, and effect size. As we assessed the risk of bias of the included studies after data collection, we extracted data related to risk assessment along with the above-mentioned data. We used Newcastle-Ottawa Scale^11^ for risk assessment because the included studies were cross-sectional and cohort studies.

#### Data synthesis

First, we contacted the corresponding author of eligible articles by email and asked for the original data if unavailable on the article or online. If we did not hear from them two weeks after sending the email, we excluded the article from the data synthesis.

If sufficient eligible data were obtained from more than one studies, we attempted to convert the data and synthesize them to obtain pooled estimates of the association between calcium intake and sleep quality.

The protocol for this review was registered with PROSPERO 2021, and any change made to the protocol was recorded on the same website. (CRD42021249642, available at: (https://www.crd.york.ac.uk/prospero/display_record.php?ID=CRD42021249642)

## 3. Results

### 3.1. Search results and the characteristics of the included studies

The initial search of the main databases yielded 2435 records. Removing duplicates (n = 630), we identified 1805 records (as of March 2024). After screening titles and abstracts, we excluded 1743 records. Then, we obtained the full texts of the remaining 62 records. Screening of the full texts excluded 42 reports due to unmatched study objectives, six reports due to unmatched study designs, six reports meeting the exclusion criteria, one report due to concerns about the integrity of statistical analysis methods and presentation of results. Consequently, the main database search retrieved seven reports^12–18^ for further examination. Thereafter, as supplementary search, we screened the first 300 records that appeared in the search results of Google Scholar and the reference lists of the seven reports identified in the main database search, and decided to include two reports^19, 20^ (figure 1).

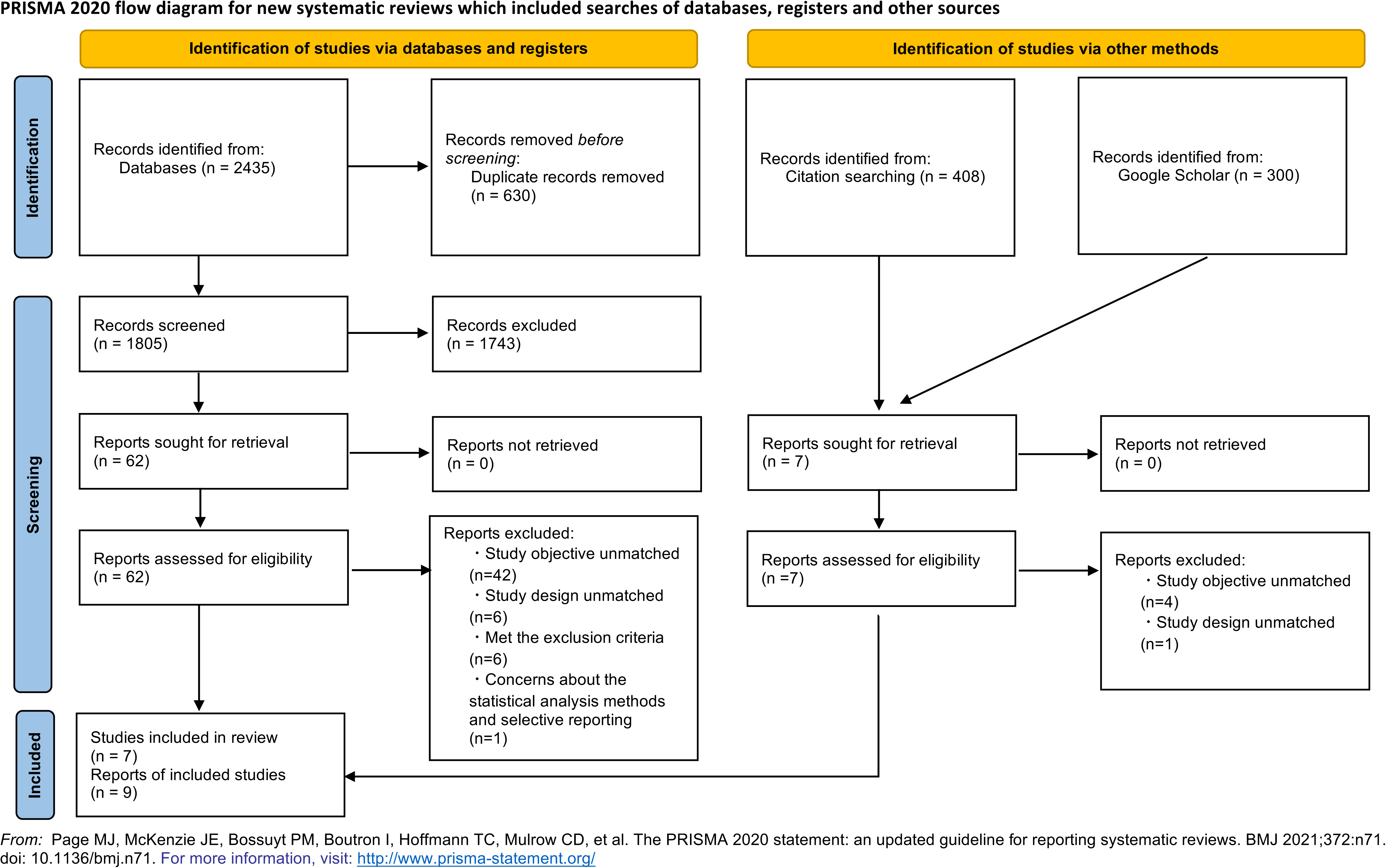

Multiple publications were identified for some of the included studies. Among the nine reports, two^12, 13^ appeared to refer to the same study according to the study settings, participants, data collection methods, and study period. The data source, target year, authors, and exposure variables were the same and the outcome variables were very similar (sleep quality and sleep duration), so we suspected that they were from one study. Three reports^15, 19, 20^ focused on secondary data analyses of the same data source (large survey datasets from the US). Two of them^15, 19^ seemed to be the same study but presented in two reports, as the authors, data source, variables and study objectives were all the same like the above case. Only the outcome variable slightly differed from that of the other report, and so we judged they were published separately but from the same study. Whereas, one report^20^ was considered a different study based on the authors, study objectives and variables though using the same large national datasets. As a result, we included seven independent studies presented in nine reports in the quality analysis.^12–20^

The basic characteristics of the included reports are listed (table 1). All of them were observational studies, and six of them were cross-sectional studies. The rest was a community-based cohort study. All of them were published in English. Two studies presented in three reports used sleep quality as an outcome.^12, 13, 15^ Five studies investigated the association between calcium intake and sleep duration (hours).^14, 17–20^ One study had both sleep duration and quality as outcomes.^16^

| study ID | country | exposure | outcome | sample size | effect measure and effect size |
| --- | --- | --- | --- | --- | --- |
| Alkhatatbeh 2021a[12] | Jordan | dietary calcium intake | sleep quality (Insomnia Severity Index) | 1000 | $r=-0.1$ ($p<0.01$) |
| Alkhatatbeh 2021b[13] | Jordan | dietary calcium intake | sleep quality (Pittsburgh Sleep Quality Index) | 1422 | $r=-0.11$ ($p<0.001$) |
| Mossavar-Rahmani 2015[14] | USA | dietary calcium intake | sleep duration | 11888 | adjusted mean calcium intake (mg per 1000kcal, reference group: intermediate sleepers)<br>short sleepers: -13.4 ($p<0.01$)<br>long sleepers: -5.3 ($p<0.05$) |
| Grandner 2014[15] | USA | dietary calcium intake | sleep quality | 4548 | • OR for those with a 100% increase in intake of calcium to have difficulty falling asleep: 0.78 (95%CI: 0.66-0.92)<br>• OR for those with a 100% increase in intake of calcium to have difficulty maintaining sleep: 0.82 (95%CI: 0.70-0.95)<br>• OR for those with a 100% increase in intake of calcium to have non-restorative sleep: 0.9 (95%CI: 0.77-1.06)<br>• OR for those with a 100% increase in intake of calcium to have daytime sleepiness: 0.89 (95%CI: 0.76-1.04) |
| Cakir 2019[16] | Turkey | dietary calcium intake | sleep quality (Pittsburgh Sleep Quality Index)<br>sleep duration | 2446 | average calcium intake (mg, reference group: good sleep quality)<br>poor sleep quality: -23.84, $p=0.037$<br>average calcium intake (mg, reference group: sleep duration 6-8 h)<br>sleep duration <6h: -6.5<br>sleep duration >8h: -23.1<br>$p=0.173$ |
| Komada 2017[17] | Japan | dietary calcium intake (mg/1000 kcal) | sleep duration | 1907 | correlation coefficient between calcium intake and sleep duration<br>men: 0.218, women: 0.208 |
| Nguyen 2023[18] | Republic of Korea | dietary calcium intake | sleep duration | 2084 | average calcium intake (mg, reference group: sleep duration 7-8h)<br>sleep duration <5h: -25.65<br>sleep duration 5-6h: 27.04<br>sleep duration >9h: -26.19<br>adjusted $p=0.002$ |
| Grandner 2013[19] | USA | dietary calcium intake | sleep duration | 4548 | difference in calcium intake (mg, reference group: normal sleep duration)<br>very short: -65<br>short: -33<br>long sleep duration: -88<br>$p=0.002$ |
| Zhu 2021[20] | USA | dietary calcium intake | sleep duration | 1783 | adjusted RR for sleep duration (calcium intake +1SD, relative to 7-8h)<br>• pre/peri-menopausal<br><5h sleep: 0.46 ($p<0.1$)<br>5-6h sleep: 1.18<br>>=9h sleep: 0.88<br>• post-menopausal<br><5h sleep: 0.62<br>5-6h sleep: 0.68<br>>=9h sleep: 0.5 |

r: correlation coefficient, OR: odds ratio, RR: risk ratio

### 3.2. Risk of bias in the included studies

Of the nine reports, three scored 5 out of 10 points,^12, 13, 16^ five scored 8 points,^14, 15, 18–20^ and the rest scored 6 points (figure 2).^17^

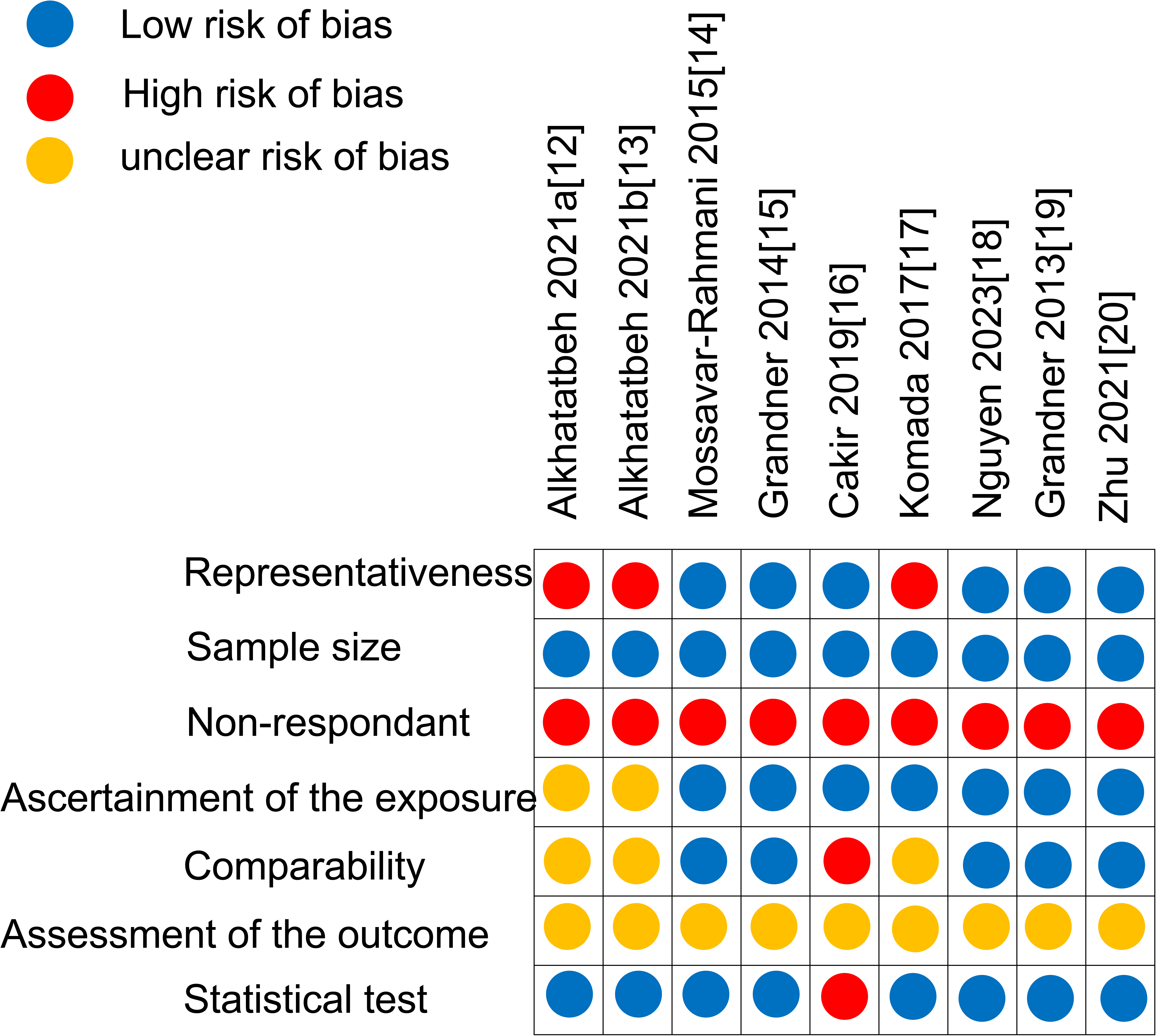

Most of the included studies were evaluated as good, for sample size and statistical test. On the other hand, they did not have adequate response rate, nor description of the response rate or the characteristics of the responders and the non-responders. All of them used self-report for the outcome assessment (sleep quality or sleep duration), which is less reliable than blinded objective assessments.

As for representativeness of the sample, ascertainment of the exposure (calcium intake) and comparability, the results varied across reports. We suspected that studies participants of which were recruited by a company were at higher risk of selection bias,^17^ as well as the studies focusing mainly on young adults.^12, 13^ In order to assess the exposure (calcium intake), five studies used 24-hour recall.^14–16, 18–20^ One study used brief self-administered diet history questionnaire.^17^ Both have been validated as assessment tools of dietary intake. However, we considered that a self-administered questionnaire used in one study (presented in two reports) might have not been valid because it had been used only in one previous study conducted by the same authors.^12, 13^ They did not mention the validity of the questionnaire in the reports.

Regarding comparability, some studies scored low for not controlling the statistical analyses for important confounding factors. Some studies considered the use of calcium supplementation and conducted regression analyses, but others did not mention confounding factors or calcium supplementation.

### 3.3. Association between calcium intake and the outcomes

Some studies did not clearly define the exposure and the outcome. In addition, it appeared that sleep quality/duration was an exposure and calcium intake was an outcome in the objectives or main analyses of some of the included studies. In these cases, we assumed that they meant calcium intake as an exposure and sleep quality as an outcome.

For exposure assessment (calcium intake), five studies presented in six reports^14–16, 18–20^ used 24-hour recall. One study^17^ used brief self-administered diet history questionnaire (BDHQ). In one study presented in two reports,^12, 13^ calcium intake was calculated based on the daily frequency and type of dairy products consumed, including cheese, yogurt, milk and labanah.

### Calcium intake and sleep quality

Of the three studies that investigated the association between calcium intake and sleep quality, one used the frequency of sleep problems^15^ and the other used the Pittsburgh Sleep Quality Index (PSQI)^16^ to assess sleep quality. One study presented in two reports used Insomnia Severity Index (ISI) and PSQI (table 1).^12, 13^

Alkhatatbeh et al., whose study was presented in two reports, used the correlation coefficient^12, 13^ between ISI/PSQI scores and calcium intake (mg/day) for the effect measure. They found a positive correlation between calcium intake and sleep quality. Participants presenting no clinically significant insomnia reported their calcium intake of 310.1 mg/day (median). Participants presenting subthreshold insomnia reported 277.3 mg/day (median), and those presenting moderate to severe clinical insomnia reported 219.6 mg/day (median).^12^ Participants presenting good sleep quality reported their calcium intake of 348.9 mg/day (mean), while participant presenting poor sleep quality reported 260.7 mg/day (mean).^13^

Grandner et al. used odds ratio (OR) for the association between dietary calcium intake and sleep-related difficulties.^15^ They found that increased calcium intake was associated with decreased difficulty falling asleep and non-restorative sleep.^15^ The OR for those with a 100% increase in calcium intake to have difficulty falling asleep was 0.78 (95%CI: 0.66-0.92). The OR for those with a 100% increase in calcium intake to have difficulty maintaining sleep was 0.82 (95%CI: 0.70-0.95). The OR for those with a 100% increase in calcium intake to have non-restorative sleep was 0.9 (95%CI: 0.77-1.06). The OR for those with a 100% increase in calcium intake to have daytime sleepiness was 0.89 (95%CI: 0.76-1.04).

Cakir et al. used the difference in the mean calcium intake between groups with different levels of sleep quality.^16^ They found that average calcium intake was higher among those with good sleep quality than those with poor sleep quality.^16^ Participants presenting good sleep quality reported their calcium intake of 640.7 mg/day (mean), while participant presenting poor sleep quality reported 616.9 mg/day (mean).^16^

### Calcium intake and sleep duration

Of the seven studies that investigated the association between calcium intake and sleep duration,^14, 16–20^ each study used different thresholds to categorize sleep duration into groups.

Four studies used difference in mean calcium intake between groups with different sleep duration categories for effect measure.^14, 16, 18, 19^ Mossavar-Rahmani et al. found that calcium intake of short sleepers and long sleepers was significantly lower than that of intermediate sleepers.^14^ Participants presenting short sleep duration reported their calcium intake of 408.8 mg/day (adjusted mean), and participant presenting long sleep duration reported 416.9 mg/day (adjusted mean), while participants presenting intermediate sleep reported 422.2 mg/day (adjusted mean).^14^ Cakir et al. found that calcium intake among those with sleep duration of less than 6 hours or more than 8 hours was lower than those with sleep duration of 6 to 8 hours, but the difference appeared not statistically significant (p=0.173).^16^ Participants presenting short sleep duration reported their calcium intake of 632.9 mg/day (mean), and participant presenting long sleep duration reported 616.3 mg/day (mean), while participants presenting intermediate sleep duration reported 639.4 mg/day (mean).^16^ Nguyen et al. found that average calcium intake was lower among people with short or long sleep duration (<5 hours: 370.6 mg/day [median], >9 hours: 370.1 mg/day [median]) than those with adequate sleep duration (5-6 hours: 423.4 mg/day [median], 7-8 hours: 396.3 mg/day [median]).^18^ Participants presenting very short sleep duration reported their calcium intake of 894 mg/day (mean), participants presenting short sleep duration reported 926 mg/day (mean), and participant presenting long sleep duration reported 871 mg/day (mean), while participants presenting normal sleep duration reported 959 mg/day (mean).^19^

Komada et al.^17^ found that there was no significant correlation (p>0.05) between dietary calcium intake and sleep duration among both men and women.^17^

Zhu et al.^20^ found that risk of short or long sleep hours (<5 hours or >9 hours compared to 7-8 hours) decreases with increasing calcium intake in pre/peri-menopausal women.^20^

#### Meta-analysis and assessment of publication bias

After examining the feasibility of meta-analysis, we decided not to conduct one because of the insufficient number of studies. We sent emails to corresponding authors of six studies to ask for the original data, and three of them replied. Among these three studies, two used data from the National Health and Nutrition Examination Survey in the United States. We concluded that these two studies were not eligible for meta-analysis as they were secondary data analyses and the national survey datasets were too large to merge with other data.

We also attempted to merge the average calcium intake data for each category of the outcome variables (sleep quality and sleep duration) from the included studies. However, one study presented in two reports^12, 13^ that addressed sleep quality as an outcome variable appeared to use the same datasets as mentioned above. Therefore, we agreed that there was no point in merging the results from these studies as the original data of them are the same. In addition, two studies^13, 16^ used PSQI scores, but the thresholds for categorization of participants into good or poor sleep quality were different between these studies. We were not able to re-categorize or convert the data from these two studies because the original data were not available.

Likewise, all studies addressing sleep duration as an outcome variable used different thresholds for categorization of sleep duration (e.g., short, good, or long). Therefore, we were not able to merge the results of these studies to obtain pooled estimates of the association between calcium intake and sleep duration.

In addition, we did not assess publication bias using a funnel plot because we did not find more than 10 studies with the same types of exposure and outcome measures.

## 4. Discussion

In this systematic review, we examined the existing evidence for the association between calcium intake and sleep quality/duration. Most of the included studies presented that calcium intake was associated with sleep quality though the results from some reports were not statistically significant. As for sleep duration, most included studies showed statistical significance for its association with calcium intake.

In this review, most included studies did not refer to confounder, and it is still unclear if the association between calcium intake and sleep quality is altered by confounders or effect modifiers. One example is vitamin D. Active vitamin D plays an important role in the metabolism and absorption of calcium gained from food.^21^ A previous systematic review shows that vitamin D deficiency can increase the risk of sleep disorders including poor sleep quality, short sleep duration, and sleepiness.^22^ This finding implies that vitamin D may be highly involved in the calcium-sleep association.

Despite such findings, the association between calcium intake and sleep quality/duration might be confounded by several factors, which none of the included studies considered. For example, people who do not pay attention to food may have no interest in healthy lifestyle, including sleep durations, indicating that attention to a healthy lifestyle could be a confounder.

This systematic review has several strengths. First, we searched for the best available databases without restricting publication years and language as long as the title and abstract were written in English, as well as screening the reference lists of the included reports. Second, to minimize information bias in this systematic review, we assessed the risk of bias in the included articles. Two reviewers (AI and JK) independently assessed the quality of included studies and double-checked the extracted data. Search results (records) were also screened by the two reviewers independently to minimize selection bias in this review. We contacted the corresponding authors of included studies for raw data to avoid missing potentially relevant data.

This systematic review, however, has some limitations. First, there were insufficient studies to conduct a meta-analysis and we could not obtain pooled estimates for the association between calcium intake and sleep quality/duration. Second, the included studies did not use the same tools to assess the calcium intake and the units of intake measure differed across studies. The assessment of sleep quality also varied across studies. The outcome of this systematic review, sleep quality, is subjective, and the results can vary depending on the assessment method. Although validated assessment tools which objectively measure sleep quality are available, such as PSQI,^23^ we found few studies that used validated assessment tools. Therefore, it was difficult to compare or merge the results of the included studies. There are also concerns that some studies did not consider confounders, such as sex, age, and ethnicity, which can affect the results. Finally, the quality of most of the included studies was not sufficiently high with a high risk of bias, which affects the overall result of this systematic review.

In conclusion, we found evidence of a positive relationship between calcium intake and sleep quality and sleep duration. To improve the sleep quality of people, we suggest promoting eating more food containing calcium in countries where the average calcium intake is low, such as Japan and South Korea. To encourage the consumption of calcium-rich food, educational programs may be useful to lead to higher quality of sleep. In clinical practice, it can be effective to assess the daily calcium intake of patients with sleep problems and prescribe calcium supplements (e.g., tablets) to improve their sleep quality. However, patients with other health problems such as mental disorders, may not be eligible for this program.

Finally, current evidence is limited. Included studies were all observational studies, and no intervention study has been conducted on this research topic. From this systematic review, it also became evident that few high-quality studies with minimal bias have been conducted. To test the effectiveness of calcium intake on sleep quality, a randomized controlled trial is required using validated measurement tools for sleep quality, such as the PSQI for outcome assessment. Moreover, studies are expected to examine the relationship between calcium intake and sleep quality in different geographical areas as diet or sleep patterns may differ depending on the culture in different regions.

## Data Availability

All data produced are available online.

---

**Appendix 1:**
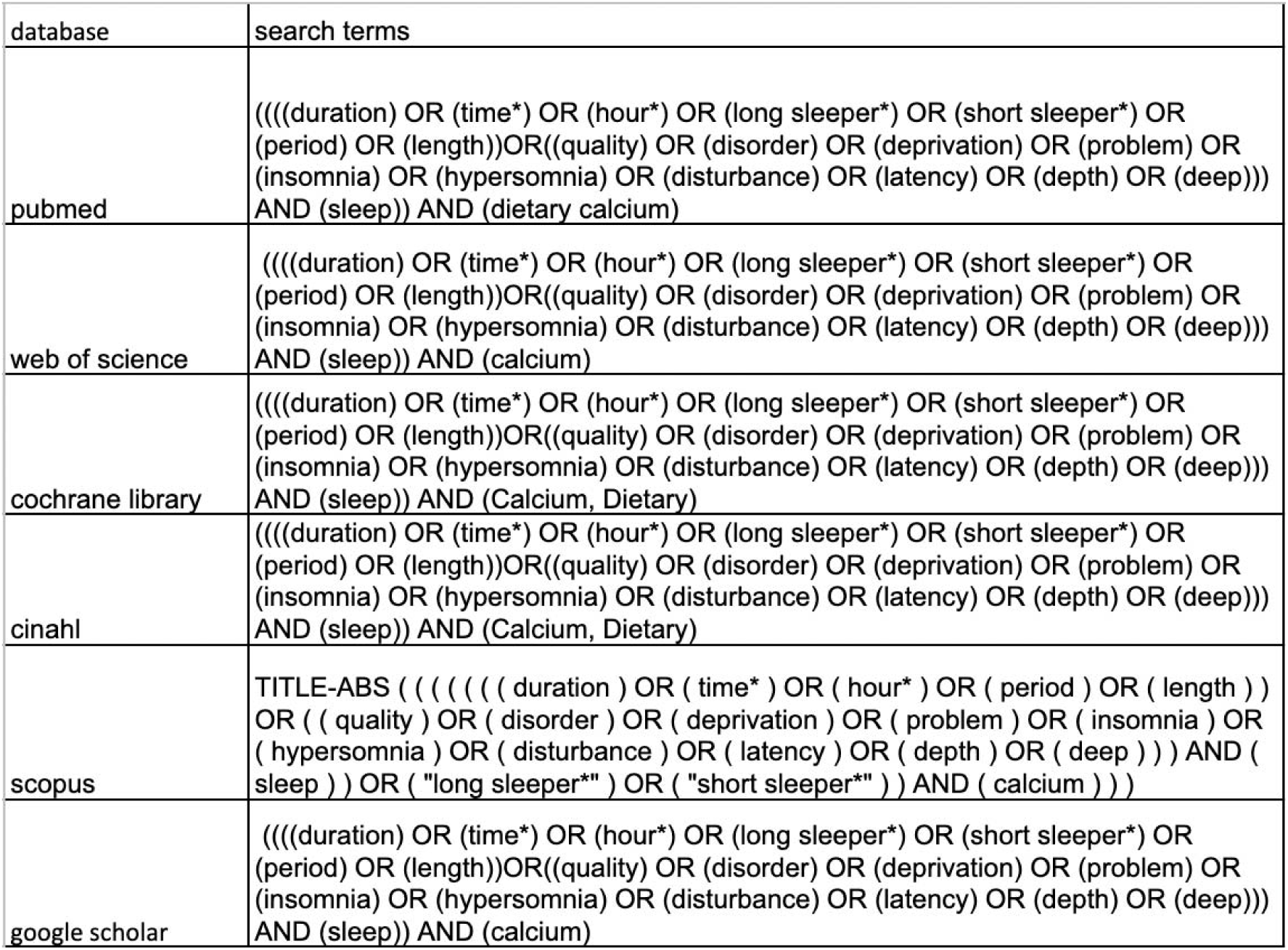
Search strategies.

